# Sensitivity of SARS-CoV-2 antibody tests with late convalescent sera

**DOI:** 10.1101/2021.07.07.21259772

**Authors:** Judith Kannenberg, Carolin Schnurra, Nina Reiners, Reinhard Henschler, Raymund Buhmann, Thorsten Kaiser, Ronald Biemann, Mario Hönemann, Grit Ackermann, Henning Trawinski, Christian Jassoy

**Author notes:** Corresponding author: Christian Jassoy, M. D., Institute for Medical Microbiology and Virology, University Clinics, University of Leipzig, Johannisallee 30, 04103 Leipzig.

## Abstract

SARS-CoV-2-specific IgM antibodies wane during the first three months after infection and IgG antibody levels decline. This may limit the ability of antibody tests to identify previous SARS CoV-2 infection at later time points. To examine if the sensitivity of antibody tests falls off, we compared the sensitivity of two nucleoprotein-based antibody tests, the Roche Elecsis II Anti-SARS-CoV-2 and the Abbott SARS-CoV-2 IgG assay and three glycoprotein-based tests, the Abbott SARS-CoV-2 IgG II Quant, Siemens Atellica IM COV2T and Euroimmun SARS-CoV-2 assay with 56 sera obtained 6-8 months after SARS-CoV-2 infection. The sensitivity of the Roche, Abbott SARS-CoV-2 IgG II Quant and Siemens antibody assays was 94.6 % (95% confidence interval (CI) 85.1-98.9 %), 98.2 % (95% CI: 90.4-99.9 %) and 100 % (95% CI: 93.6-100 %). The sensitivity of the N-based Abbott SARS-CoV-2 IgG and the glycoprotein-based Euroimmun ELISA was 48.2 % (95% CI: 34.7-62.0 %) and 83.9 % (95% CI: 71.7-92.4 %). The nucleoprotein-based Roche and the glycoprotein-based Abbott RBD and Siemens tests were more sensitive than the N-based Abbott and the Euroimmun antibody tests (p=0.0001 to p=0.039). The N-based Abbott antibody test was less sensitive 6-8 months than 4-10 weeks after SARS-CoV-2 infection (p = 0.0002). The findings show that most SARS CoV-2 antibody assays correctly identified previous infection 6-8 months after infection. The sensitivity of pan-Ig antibody tests was not reduced at 6-8 months when IgM antibodies have usually disappeared. However, one of the nucleoprotein-based antibody tests significantly lost sensitivity over time.

**Highlights:**

- Most antibody tests correctly identified SARS CoV-2 infection 6-8 months after infection
- The sensitivity of the antibody tests was 48.2-100 %
- The three tests with the highest sensitivity (94.6-100 %) were the N-based Roche and the RBD-based Abbott and Siemens assays
- The N-based Abbott IgG CMIA was significantly less sensitive 6-8 months than 4-10 weeks after infection (p = 0.0002)

## 1. Introduction

Coronavirus disease 2019 (COVID-19) caused by the severe acute respiratory syndrome coronavirus 2 (SARS-CoV-2) has provoked a global pandemic. As of February 13, 2021, 107,686,655 people have been infected globally and 2,368,571 deaths have been reported to WHO.

SARS-CoV-2 antibody tests help in determining seroprevalence and identifying previously infected individuals. They are useful in symptomatic patients with repeatedly negative nucleic acid amplification test and in children with multisystem inflammatory syndrome [1–3]. Numerous studies have examined the sensitivity and specificity of available antibody tests. As the epidemic started only recently, the studies used sera that were obtained early after infection. Few data exist about the performance of antibody tests in individuals more than six months after infection [2–9].

SARS-CoV-2 antibody tests are either being performed at point of care or in the diagnostic laboratory. Laboratory SARS-CoV-2 antibody tests either detect IgG, IgM, IgA, IgG plus IgM or all antibody classes. In the early postinfection period, most sera contain virus-specific IgM, IgG and IgA [10,11]. The proportion of sera with IgM antibodies reaches a peak at 4-5 weeks and the percentage of positive sera subsequently declines [12]. It was reported that the combined measurement of SARS-CoV-2-specific IgG and IgM antibodies is more sensitive than measurement of either antibody alone [13]. This raised the question if the sensitivity of antibody tests for all antibody classes declines in late convalescence when IgM antibodies have disappeared.

SARS-CoV-2 antibody assays measure antibodies against the viral nucleoprotein (N), the glycoprotein spike 1 (S1), the glycoprotein spike 2 (S2), the receptor binding domain of S1 (RBD), or a combination of several viral proteins. As a group, SARS CoV-2 nucleoprotein- and glycoprotein-based antibody tests showed similar sensitivity when tested with sera from the early weeks to months after infection [3,9,14]. It was reported that the SARS-CoV-2 nucleoprotein-specific antibody response decays with a half-life of 55-90 days and the RBD-specific antibodies with a T1/2 of 66-235 days [15]. It was also observed that the percentage of antibody positive individuals declines over time [15,16]. This suggests that over time SARS CoV-2 antibody tests lose the ability to identify previously infected individuals.

The aim of the study was to determine the ability of five antibody immunoassays to diagnose previous SARS CoV-2 infection 6-8 months after infection. The tests reflected different technical approaches of SARS-CoV-2 antibody testing. They were based on enzyme immunoassay (ELISA), chemiluminescence microparticle immunoassay (CMIA), microparticle immunoassay (MIA) or electrochemiluminescence immunoassay (ECLIA) technology, detected either all antibody classes or IgG and targeted antibodies against either the viral nucleoprotein or the glycoprotein.

## 2. Study design

A prospective diagnostic study was performed to examine the sensitivity of five commercial SARS-CoV-2 antibody tests with late convalescent sera.

### 2.1 Serum samples

A total of 56 venous blood samples were obtained from 55 adults 6-8 months after recovery from COVID-19 (53 at 6, two at 7, one at 8 months). The majority of the patients had mild (headache, common cold, cough) to moderate (fever, myalgia, abnormal fatigue) symptoms, some were asymptomatic and none of them required hospitalization. The participants (except CoV-046) had previously participated in a study about the diagnostic sensitivity of SARS-CoV-2 antibody tests 2-10 weeks after the infection [14]. The study was approved by the Local Ethics Commission at the Medical Faculty at the University of Leipzig (ethical vote 147/20-ek). Sera were obtained after informed consent. All participants except one (CoV-046) had a positive PCR test. CoV-046 was diagnosed clinically and showed a positive result in all antibody tests (Suppl. Table 1). Sera were stored at −20 °C until testing.

### 2.2 Antibody tests

Sera were analyzed with three tests that measure antigen-specific IgG and two assays for all immunoglobulin classes. The tests were specific for antibodies against either the viral nucleoprotein or parts of the glycoprotein. The Roche Elecsys Anti-SARS-CoV-2 is a bridging ruthenium complex ECLIA for nucleoprotein-specific antibodies of all classes (IgG, IgM, other Ig). It was performed with an automated cobas e 601 analyzer. The Abbott SARS-CoV-2 IgG and SARS-CoV-2 IgG II Quant assays are acridinium CMIA for the detection of IgG antibodies against the nucleoprotein (SARS-CoV-2 IgG) or glycoprotein receptor binding domain (SARS-CoV-2 IgG II Quant). The assays were performed with the ARCHITECT i2000SR system. The Siemens Atellica IM COV2T is a bridging acridiniumester chemiluminescence MIA intended to detect IgG, IgM and other immunoglobulins against the receptor-binding domain (RBD) of S1 glycoprotein. It was run on the Atellica IM analyzer. The Euroimmun Anti-SARS-CoV-2 IgG ELISA measures antibodies against the S1 domain of the spike protein and was performed with an automated ELISA processor (DSX, Dynex Technologies, U.K.). The tests were conducted in three diagnostic routine laboratories.

### 2.3 Data analysis

Medcalc statistical online software was used (https://www.medcalc.org/calc/) for data analysis. Equivocal sera were counted as negative. The „Test for one proportion” based on the “exact” Clopper-Pearson confidence interval was utilized to calculate the 95% confidence intervals of test positive rates. The sensitivity of the tests at different time points was compared with the calculator „Comparison of proportions” that uses the “N-1” Chi-squared test. The sensitivity of the tests with paired serum samples was compared with a mid-p McNemar test using R statistical software. A significant difference was defined as p < 0.05. Simple calculations and data visualization were performed with LibreOffice Calc.

## 3. Results

### 3.1 Sensitivity of the antibody tests

The sensitivity of the tests ranged from 48.2 % to 100 %. The N-based Roche test recognized 94.6 % of the sera (95% confidence interval (CI): 85.1-98.9 %). The N-based Abbott test was positive with 48.2 % (95% CI: 34.7-62.0 %) of the samples. The glycoprotein RBD-based Abbott test showed 98.2 % positive results (95% CI: 90.4-99.9 %). The RBD-based Siemens test was positive with all sera (100 %, 95% CI: 93.6-100 %) and the glycoprotein S1-based Euroimmun IgG antibody test showed positive results with 83.9 % (95% CI: 71.7-92.4 %) of the samples (Table 1 and Suppl. Table 1).

**Table 1:** Sensitivity of SARS-CoV-2 antibody tests with sera obtained 6-8 months after infection.

|  | **Roche** | **Abbott N** | **Abbott RBD** | **Siemens** | **Euroimmun** |
| --- | --- | --- | --- | --- | --- |
| **Antibody** | All Ig classes | IgG | IgG | All Ig classes | IgG |
| **Antigen<sup>1</sup>** | N | N | RBD | RBD | S1 |
| **Sensitivity** | 94.6 % | 48.2 % | 98.2 % | 100 % | 83.9 % |
| **95% confidence interval** | 85.1-98.9 % | 34.7-62.0 % | 90.4-99.9 % | 93.6-100 % | 71.7-92.4 % |

<sup>1</sup>N: Nucleoprotein, S1: glycoprotein spike 1 fraction, RBD: glycoprotein receptor binding domain

Among the N-based tests, the Roche test that measures all antibody classes was more sensitive than the IgG-specific Abbott test (p < 0.0001). Among the glycoprotein-based tests the Abbott SARS-CoV-2 IgG II Quant and the Siemens MIA showed comparable sensitivity (p = 0.5) and the two tests were more sensitive than the Euroimmun IgG ELISA (p = 0.0039 and 0.0019). Comparison of N- and glycoprotein-based antibody tests showed that the N-based Roche assay was more sensitive than the glycoprotein-based Euroimmun assay (p = 0.039). The Roche assay showed fewer positive results than the RBD-based Abbott and Siemens tests (53 versus 55 and 56 positive sera), but the differences were not statistically significant (p = 0.375 and 0.125) (Table 2).

**Table 2:** Significance level (p-value) for difference of test sensitivity^1^.

| **Test** | **Abbott N** | **Abbott RBD** | **Siemens** | **Euroimmun** |
| --- | --- | --- | --- | --- |
| **Roche** | < 0.0001* | 0.375 | 0.125 | 0.039* |
| **Abbott N** | x | < 0.0001* | < 0.0001* | < 0.0001* |
| **Abbott RBD** | x | x | 0.5 | 0.0039* |
| **Siemens** | x | x | x | 0.0019* |

<sup>1</sup> The tests were compared with the mid-p McNemar test
\* Significant difference ( $p < 0.05$ ) is being labeled with an asterisk.

### 3.2 Comparison of positive rates of early convalescent and late convalescent sera

We compared the percentage of positive results with data obtained with sera from mostly the same individuals 4-10 weeks after infection [14]. Comparison of the positive rates of the antibody tests with early and late convalescent sera showed that the N-based Abbott test was markedly less sensitive 6-8 months than 4-10 weeks after infection (p = 0.0002). The sensitivity of the Roche, Siemens, and Euroimmun tests was similar 4-10 weeks and 6-8 months after infection (Table 3).

**Table 3:** Percentage of positive results and significance level of differences of antibody positivity rates at 4-10 weeks and 6-8 months after infection^1^.

|  | **Number of sera** | **Roche** | **Abbott N** | **Siemens** | **Euroimmun** |
| --- | --- | --- | --- | --- | --- |
| **4-10 weeks** | 48 | 95.8 % | 83.3 % | 100 % | 89.6 % |
| **6-8 months** | 56 | 94.6 % | 48.2 % | 100 % | 83.9 % |
| **Difference** | - | 1.2 % | 35.1 % | 0 % | 5.7 % |
| **Significance level (p)** | - | 0.7784 | 0.0002* | - | 0.4023 |

<sup>1</sup> The tests were compared with the Chi-squared test
\* Significant difference ( $p < 0.05$ )

## 4. Discussion

In the first 8 months after infection, the antibody response to SARS CoV-2 declines with half-lifes of 55-90 and 66-235 days for nucleoprotein- and RBD-specific antibodies, respectively [15]. This suggests that antibody tests may lose the ability to identify previous infection. Moreover, as the half-life of nucleoprotein-specific antibodies was reported to be shorter, tests that measure antibodies against the viral nucleoprotein may be affected to a larger degree. Similarly, some of the antibody tests are specific for single Ig classes such as IgG, some tests detect all antibody classes. These tests may lose sensitivity at time points when IgM responses have usually disappeared.

Three of the antibody tests had sensitivities of 94.6-100 %. One of the tests was specific for antibodies against the viral nucleoprotein and two tests measured antibodies against RBD. This shows that in principle, antibody tests specific for antibodies against both viral proteins are appropriate for testing at 6-8 months after infection. The RBD-based antibody tests recognized slightly more sera than the N-based test, but this was not statistically significant and needs to be examined with more serum samples.

It was previously reported that the combined measurement of SARS-CoV-2-specific IgG and IgM antibodies is more sensitive than measurement of either antibody alone [13]. It was also observed that after SARS-CoV-2 infection the proportion of patient sera that are positive for virus-specific IgM declines from more than 90 % early after infection to 22.7-30.8 % 3-6 months after symptom onset [12,17]. This raised the question whether the sensitivity of SARS-CoV-2 antibody tests for all antibody classes decreases to levels similar to that of antibody tests for virus-specific IgG when testing sera from later time points. Two of the three highly sensitive tests were bridging assays that measure antibodies of all classes. One of the tests was IgG-specific. Thus, the pan-Ig antibody assays were highly sensitive independent of the presence of IgM.

Comparison of the percentage of positive samples at 4-10 weeks and 6-8 months after infection showed that the sensitivity of three of the tests was similar at the two time points. We did not compare the sensitivity of the Abbott RBD-based antibody assay, but a decline in sensitivity is unlikely because of the sensitivity of 98.2 % at 6-8 months. Thus, four of the five tests showed high sensitivity for 6-8 months after infection.

The percentage of positive results of the N-based Abbott assays was significantly lower than that of the other assays. Similarly, the positivity rate declined significantly 6-8 months compared with 4-10 weeks after infection. At 6-8 months after infection, less than half of the sera was positive. The numbers are similar to those reported by Lumley et al. who found that the median time remaining nucleocapsid IgG positive with the N-based Abbott assay was 166 days [18]. A possible explanation for the limited sensitivity of this test at 6-8 months is the faster decline of N-specific than glycoprotein-specific antibodies as reported previously [15]. In addition, the antibody assay may have been designed for maximum specificity rather than sensitivity. This is beneficial at low seroprevalence to maximize the positive predictive value [3,14]. The low sensitivity of the N-based Abbott IgG antibody assay indicates that the optimal use of the test is in the period shortly after presumed infection.

The study shows that, in principle, various technical antibody test platforms including ELISA, CMIA, bridging ECLIA and bridging MIA were appropriate [7,13,14]. The study did not test N-based ELISAs or point-of-care tests. In addition, the study did not examine sera beyond 8 months after infection. Presumably, the percentage of positive sera will further decline. It needs to be examined if the more rapid decline of N-specific antibodies reduces the sensitivity of N-based tests disproportionally. The serum panel included specimens from participants with asymptomatic, mild or moderate disease. As more severe infections lead to more robust immune responses, the sensitivity of SARS-CoV-2 antibody tests would possibly be higher with sera from such individuals.

In summary, our findings show that the N-based Roche and RBD-based Abbott and Siemens SARS-CoV-2 antibody immunoassays as well as the glycoprotein S1-based Euroimmun IgG antibody ELISA were highly sensitive 6-8 months after SARS-CoV-2 infection. The decline of IgM in the first three months did not affect the performance of the pan-Ig assays. These antibody tests are appropriate for diagnostic and epidemiological purposes when the time point of infection may date back more than a couple of months. The N-based Abbott antibody test showed low sensitivity at 6-8 months and significantly lost sensitivity compared with sera from 4-10 weeks indicating that this test is optimal early after infection.

## Data Availability

The data referred to in the manuscript will be made accessible upon request

## Declaration of Competing Interest

The authors declare that there are no conflicts of interests.

## Acknowledgement

We thank the participants of the study for participating, Dr. Elvira Edel and the clinical research team at the Institute for Transfusion Medicine for blood drawings and Dr. Maciej Rosolowski for helpful advice with the statistics. The study was supported by the Faculty of Medicine at the University of Leipzig and funded in part by grant no. 1-0421/31/40-2020/26882 from the Free State of Saxony and grant no. JA 662/22-1 from the Deutsche Forschungsgemeinschaft.

## ^1^Abbreviations

SARS-CoV-2: Severe acute respiratory syndrome coronavirus-2
COVID-19: Corona-virus disease 2019
N: nucleoprotein
RBD: receptor binding domain
ELISA: enzyme immunoassay
CMIA: chemiluminescence microparticle immuno-assay
MIA: microparticle immunoassay
ECLIA: electrochemiluminescence immunoassay

## Credit authorship contribution statement

Judith Kannenberg: Methodology, formal analysis, investigation, drafting the manuscript, review and editing. Carolin Schnurra: Methodology, recruitment of participants, storage of samples, writing - review and editing. Nina Reiners: Recruitment of participants, storage of samples. Reinhard Henschler: Resources, writing-review and editing. Raymund Buhmann: Organization of blood drawing. Thorsten Kaiser: Acquisition of data. Ronald Biemann: Acquisition of data, writing – review & editing. Mario Hönemann: Acquisition of data, writing – review and editing. Grit Ackermann: Acquisition of data. Henning Trawinski: Methodology, recruitment of participants, writing - review and editing. Christian Jassoy: Conceptualization, methodology, formal analysis, writing – review and editing.

## Notes

### Competing Interest Statement

The authors have declared no competing interest.

### Author Declarations

The study was approved by the Local Ethics Commission at the Medical Faculty at the University of Leipzig (ethical vote 147/20-ek)

