## Supplemental Table 1 for "Sensitivity of SARS-CoV-2 antibody tests with late convalescent sera"

**Supplementary Table S1**: Comparison of the reactivity of SARS-CoV-2 antibody tests

| Assay | Roche | Abbott N | Abbott RBD | Siemens | Euroimmun |
|---|---|---|---|---|---|
| Test units | N-COI | N-Index | AU/ml | S-Index | S-Ratio |
| Cut-off* | 1,00 | <0.5 neg.,≥1.4 pos. | 50.0 AU/ml | 1,00 | <0.8 neg., ≥1.1 pos. |
| CoV-030-2** | 237,80 | 6.13 | 487.9 | > 10.00 | 4,47 |
| CoV-013-2 | 193,10 | 5.35 | 2877.2 | > 10.00 | 9,72 |
| CV220/006-3 | 194,20 | 5.31 | 589.1 | > 10.00 | 4,04 |
| CoV-035-2 | 108,50 | 3.96 | 778.3 | > 10.00 | 4,91 |
| CV220/035-3 | 227,50 | 3.63 | 549.2 | > 10.00 | 4,58 |
| CoV-022-2 | 224,80 | 3.57 | 390.0 | > 10.00 | 2,91 |
| CoV-004-2 | 234,10 | 3.48 | 2020.2 | > 10.00 | 7,18 |
| CoV-016-2 | 220,80 | 3.48 | 1015.7 | > 10.00 | 6,79 |
| CoV-029-2 | 192,70 | 3.39 | 1558.3 | > 10.00 | 8,65 |
| CoV-020-2 | 117,80 | 3.34 | 41.1 | 1,89 | 0,72 |
| CoV-031-2 | 29,72 | 2.67 | 182.2 | > 10.00 | 1,93 |
| CoV-006-2 | 139,80 | 2.51 | 603.5 | > 10.00 | 3,52 |
| CV220/035-4 | 178,40 | 2.50 | 534.5 | > 10.00 | 4,36 |
| CV220/001-3 | 85,31 | 2.25 | 689.8 | > 10.00 | 4,94 |
| CoV-002-2 | 124,20 | 2.16 | 815.2 | > 10.00 | 6,93 |
| CoV-023-2 | 133,90 | 2.14 | 353.5 | > 10.00 | 3,45 |
| CoV-026-2 | 38,78 | 1.98 | 1550.4 | > 10.00 | 6,83 |
| CoV-046-1 | 95,50 | 1.93 | 171.4 | > 10.00 | 2,28 |
| CV220/011-5 | 77,19 | 1.85 | 432.2 | > 10.00 | 3,16 |
| CV220/010-4 | 105,70 | 1.83 | 1360.1 | > 10.00 | 8,07 |
| CoV-025-2 | 96,19 | 1.80 | 173.0 | > 10.00 | 1,51 |
| CoV-021-2 | 37,36 | 1.75 | 958.3 | > 10.00 | 6,88 |
| CoV-003-2 | 71,22 | 1.66 | 1296.7 | > 10.00 | 5,88 |
| CoV-042-2 | 95,23 | 1.62 | 811.9 | > 10.00 | 5,91 |
| CoV-027-2 | 136,80 | 1.59 | 372.8 | > 10.00 | 2,01 |
| CoV-012-2 | 38,39 | 1.57 | 65.4 | 2,90 | 0,78 |
| CV220/008-3 | 124,60 | 1.40 | 184.8 | > 10.00 | 2,41 |
| CV220/024-3 | 111,00 | 1.35 | 866.3 | > 10.00 | 5,55 |
| CoV-045-2 | 29,06 | 1.34 | 2244.0 | > 10.00 | 7,98 |
| CoV-001-2 | 76,50 | 1.23 | 613.4 | > 10.00 | 4,12 |
| CoV-039-2 | 74,76 | 1.10 | 471.3 | > 10.00 | 5,02 |
| CV220/039-3 | 22,86 | 1.06 | 143.7 | 4,17 | 1,07 |
| CoV-028-2 | 35,93 | 0.99 | 506.9 | > 10.00 | 4,34 |
| CoV-043-2 | 8,38 | 0.97 | 123.1 | > 10.00 | 1,50 |
| CoV-044-2 | 2,39 | 0.96 | 62.0 | 7,44 | 0,73 |
| CoV-032-2 | 36,85 | 0.75 | 512.0 | > 10.00 | 4,59 |
| CoV-019-2 | 11,99 | 0.73 | 356.4 | > 10.00 | 3,59 |
| CoV-038-2 | 22,26 | 0.58 | 390.2 | > 10.00 | 5,94 |
| CoV-014-2 | 25,39 | 0.57 | 565.9 | > 10.00 | 4,11 |
| CoV-015-2 | 12,46 | 0.56 | 276.9 | 8,38 | 1,09 |
| CoV-041-2 | 44,19 | 0.51 | 339.4 | > 10.00 | 2,78 |
| CoV-017-2 | 31,12 | 0.45 | 747.3 | 5,95 | 1,23 |
| CV220/022-3 | 8,67 | 0.43 | 138.2 | 6,19 | 1,30 |
| CoV-018-2 | 25,43 | 0.43 | 133.6 | 9,84 | 1,20 |
| CoV-024-2 | 7,92 | 0.34 | 1177.6 | > 10.00 | 4,42 |
| CV220/026-3 | 21,08 | 0.34 | 169.7 | > 10.00 | 2,09 |
| CV220/013-3 | 14,04 | 0.34 | 177.0 | 4,26 | 1,25 |
| CV220/002-3 | 19,07 | 0.32 | 172.9 | > 10.00 | 1,74 |
| CoV-005-2 | 12,39 | 0.25 | 994.8 | > 10.00 | 6,72 |
| CV220/003-3 | 5,80 | 0.25 | 70.9 | 3,41 | 0,82 |
| CoV-033-2 | 3,18 | 0.24 | 71.4 | 9,39 | 0,91 |
| CV220/021-3 | 3,37 | 0.12 | 146.5 | > 10.00 | 1,33 |
| CoV-036-2 | 0,26 | 0.09 | 293.8 | 5,59 | 2,92 |
| CoV-034-2 | 3,19 | 0.08 | 105.3 | > 10.00 | 1,59 |
| CoV-011-2 | 0,23 | 0.06 | 53.9 | 5,62 | 0,94 |
| CoV-040-2 | 0,63 | 0.05 | 83.4 | 2,93 | 0,65 |

* white = positive, yellow = grey zone (equivocal), red = negative, ** Sample ID
